# Exploring the behavioural determinants of adherence to prescription for acute febrile illnesses, and development of a training and communication clinical trial intervention: a description of research methods

**DOI:** 10.1101/2020.12.01.20240929

**Authors:** Philip Horgan, Olawale Salami, Mariam Otmani del Barrio, Anjana Tomar, Sarabjit S Chadha, Buddha Basnyat, Summita Udas Shakya, David Kaawa-Magiri, Heidi Hopkins, James Kapisi, Vida Kukula, Rita Baiden, Phyu Hnin Hlaing, Frank Smithuis, Adelaide Campaore, Halidou Tinto, Manu Gautam, Ashish Pathak, Manmeet Kaur, Neelam Taneja, Alok Kr. Deb, Shanta Dutta, Jan Swasthya Sahyog, Kamini Walia, Catrin E Moore, Piero Olliaro

**Author notes:** Corresponding author: Philip Horgan, Big Data Institute, Nuffield Department of Medicine, Old Road Campus, University of Oxford, Oxford, UK, OX3 7LF, (Orchid ID 0000-0002-8140-0481). Author Contributions: Contributors: All authors co-designed the study, with lead from PH in collaboration with MOB. OS and PO (FIND) obtained funding, PH wrote the first draft with inputs from MOB, CEM, OS, PO, AT, SSC, SUS, DKM, VK, PHH, AC, MK, NS; all authors contributed to the editing of the drafts.

## Abstract

**Objective:** To explore behavioural factors relating to prescription adherence and the communication of prescription adherence messages for patients with acute febrile illness, and to develop a Training & Communication (T&C) intervention to be delivered as part of a clinical trial. The clinical trial intervention package consists of improved diagnostic tools, clinical practices and the T&C package, for children, adolescents and adults presenting with fever symptoms at outpatient facilities in five LMICs.

**Design:** Content analysis of primary, qualitative data collection, informed by the Capability, Opportunity, Motivation (COM-B) theory of behaviour, the Theoretical Domains Framework (TDF) and Behaviour Change Wheel (BCW) approach.

**Setting:** Health facilities and local communities in five LMICs in Africa and Asia.

**Participants:** Health facility prescribers and local community adults.

**Intervention:** Febrile illness is a common presentation among adults and children in primary care settings, but diagnosing the cause of fever is challenging, especially in low-resource settings. Prescribers’ and patients’ behaviours underpin treatment practices, and antibiotics are the customary fallback choice for lack of better alternatives. However, in most cases antibiotics would not be required, do not cure the ongoing infection, and may have short-term (toxicity, costs) and long-term (drug resistance) untoward effects.

Trialling new approaches including point-of-care tests and diagnostic algorithms alone would provide limited information on real-life applicability if behaviours are not accounted for.

Accordingly, we designed an innovative, multiphase, mixed methods study, combining qualitative and behaviour approaches, with a quantitative two-arm, clinic based, randomised controlled trial. Qualitative and behavioural methods are used to: support the development of the Training & Communication component of the clinical trial, collect patient information on adherence, and support recommendations for future behaviour change interventions.

This paper describes the qualitative research methods used to generate the clinical trial training and communication interventions, in support of adherence to prescriptions.

*Article Summary:* Strengths and limitations of this study

- This is the first study we know of to explore the behavioural factors affecting prescription adherence and the communication of adherence messages in the LMIC study locations.
- The use of behavioural frameworks to shape the design of data gathering topic guides has the potential to illuminate the drivers for antibiotic prescription adherence, and generate the knowledge needed to support the design of effective communication interventions.
- The local nature of behavioural drivers means it is unlikely that the research findings will be generalisable and directly usable in other locations, however the process by which behavioural drivers are identified, and the process to convert to a training and communication package intervention, are applicable beyond the study sites.
- The scope of the clinical trial intervention, of which the T&C package is one component, precludes a wider behaviour change intervention which may be beneficial to improving adherence. This will be explored in a set of intervention recommendations.
- Because the T&C intervention is one component of the package of interventions for the clinical trial, we cannot rule out the influence of other intervention components (for example the use of an increased number of diagnostic tests) on prescription adherence. Similarly, due to the intervention ‘package’ approach, we cannot conclude how much the T&C package contributed to patient recovery at Day7, the primary outcome.

## Introduction

### Increases in AMR, difficulties in diagnosis of febrile illness and required behaviours

Every day around the world healthcare providers face considerable difficulties in accurately diagnosing and managing acute febrile illness, especially in low-resource settings; as clinical algorithms cannot adequately identify the aetiologies of fever, they commonly fall back on prescribing antibiotics [1]. This practice however is not without untoward consequences: ineffective treatment for viral infections and increased risks of adverse effects, some of which are life-threatening [2] and have higher costs [3]. Inappropriate prescription, together with self-treatment, are key drivers of antimicrobial resistance (AMR) through selective pressures [4] [5].

Behaviours compound these problems. Healthcare workers must use available and appropriate diagnostics and effectively communicate prescriptions to patients and caregivers. Patients and caregivers must obtain the medicines prescribed and use them as intended. Without these essential behaviours, patients will not benefit from appropriate medication, and inappropriate antibiotic use will contribute to rising AMR.

This paper describes the research methods used to improve understanding of prescription adherence behaviours and the communication of adherence messages, and the process to develop a Training and Communication package to be implemented within the Diagnostic Use Accelerator clinical trial.

Foundation for Innovative New Diagnostics’s (FIND) AMR Diagnostic Use Accelerator programme (protocol preprint DOI: <u>10.21203/rs.3.rs-35083/v1</u>) was launched at the AMR Call to Action event in Ghana on the 18th November 2018.

In the first chapter of this programme, an innovative, multiphase, mixed methods research study was implemented; in the initial phase, qualitative methods are used to explore the behavioural determinants of the communication of adherence messages by healthcare workers, and adherence to prescription by patients and caregivers.

The research findings support the design of a Training and Communication (T&C) package containing messages of prescription adherence and training for healthcare workers in the delivery of adherence messages. These communication messages are subsequently implemented as one component of a two-arm, outpatient clinic based, randomised controlled trial. The clinical trial examines the impact of an intervention consisting of improved diagnostic tools, clinical practices and training and communication (the T&C package) on acute fever case management and antibiotic prescriptions for children, adolescents and adults presenting at outpatient facilities in five LMICs.

Within the trial, the messages of adherence to prescription are communicated to patients and caregivers. During clinical follow-up, qualitative methods are used to gather data about prescription adherence. This information is quantified and recorded as part of the quantitative case record form and used in analysis of the trial endpoints, comparing intervention and control arms.

In a further, concurrent phase of research, qualitative methods are used to explore behaviour determinants on the uptake of diagnostics by healthcare workers. Finally, the Behaviour Change Wheel [6, 7] is used as a guide to formulate recommendations for future behavioural change interventions covering both adherence to prescriptions and use of diagnostics.

The trial is being conducted in five countries Uganda, Ghana, Burkina Faso, Nepal (one partner each) and India (four partner centres) (ClinicalTrials.gov Identifier: NCT04081051), under a common protocol with local adaptions. Research partners were selected following two competitive calls, for research teams globally excluding India, and for Indian research teams.

This paper describes the first of these qualitative research phases and the subsequent development of the T&C package.

### Aims and research questions

A wide range of social and contextual factors influence patients, caregivers and healthcare workers as they diagnose, prescribe, purchase or obtain, and use the prescribed medicines. To support the required behaviours (through the T&C package), researchers need to develop a deep understanding of the social and contextual pressures that act on them [8].

In this study, qualitative methods and behavioural frameworks are used to investigate the two key behaviours:

1. Adherence to prescriptions by patients and caregivers
2. Communication of adherence messages by healthcare workers

to answer the overarching research questions:

1. What are the behavioural determinants for adherence to prescriptions by patients/caregivers who present with fever symptoms at outpatient clinics in LMICS?
2. What are the behavioural determinants of communication of adherence messages by healthcare workers to patient/caregiver, who present with fever symptoms in outpatient clinics of the selected LMICs.

The T&C package consists of:

1. communication messages for patients, and caregivers (individually adapted),
2. training for healthcare workers in the delivery of these messages.

In the clinical enrolment stage of the trial, the communication messages are delivered to the participants assigned to the intervention arm, along with their prescription, following clinical assessment and use of a package of diagnostics tests.

This paper describes the core qualitative research methods and templates developed for the study, and adapted for country specific use. In addition to exploring the behaviour determinants of adherence to prescription and the communication of messages, the methods and templates describe were used to convert identified behaviour determinants from research findings into a training and communication (T&C) package to be implemented within the Diagnostic Use Accelerator clinical trial.

Local adaptions to these methods, site T&C packages and research findings will be described in future publications.

The term ‘the study’ is used below to refer to the qualitative research component to develop the T&C package, as part of the wider Dx Accelerator clinical trial.

## Materials and Methods

The description of materials and methods fall into two sections, firstly the research methods used to understand the behaviours of parents, caregivers and healthcare workers, and secondly, the process to develop a Training and Communication package from those findings.

## Research methods

### Qualitative methods

To influence the adherence behaviours and the communication of adherence messages, the T&C package must reflect a deep and nuanced understanding of the behavioural drivers of the individuals involved.

Qualitative methods, which study subjects ‘in their natural settings, attempting to make sense of, or interpret, phenomena in terms of the meanings people bring to them.’ [9] are suited to understand ‘not only the objective nature of behaviours but also subjective meanings: individuals’ own accounts of their attitudes, motivations, behavior’ [10, 11].

We combine qualitative research methods in a constructivist paradigm, with the quantitative methods of the clinical trial in a way that is different from other studies which apply qualitative methods in areas of ‘bias, efficiency, ethics, implementation, interpretation, relevance, success and validity’ [12]. Here we use qualitative methods to:

- support the development of one intervention component of the clinical trial,
- collect patient information on adherence, which is subsequently quantified and recorded in the clinical record form
- support recommendations for future behaviour change interventions

The constructivist paradigm recognises that knowledge is socially constructed by people active in the research process and that research is a product of the values of researchers and cannot be independent of them [13]. In this study social science researchers have a background in health research in the local community and health facilities, conducting research on a range of health issues. None have previously conducted analysis of behavioural determinants of adherence to prescription.

The study uses the familiar qualitative data gathering approaches of In-depth-interview and Focus Group Discussions, and Content Analysis to generate research findings.

The Content Analysis approach, which identifies themes and patterns by subjectively interpreting the content of text data through the ‘systematic classification process of coding’, from a ‘predominately naturalistic paradigm’ [14] is suited to ‘systematically describe the meaning’ of the research materials [15]. In contrast to analytical methods that use predetermined code frames, the coding frame generated from participant responses is more flexible to code the nuance of meaning that may vary by locality.

As noted by others, behavioural determinants can be identified, and targeted [16] in the clinical trial. In this study, the intervention is limited to targeting adherence to prescription behaviours through communication messages, and to communication of adherence messages, through training of healthcare workers.

### The Behaviour Change Wheel

The Behaviour Change Wheel (BCW) guide [6, 7], incorporating the Capability, Opportunity, Motivation, Behaviour (COM-B), and Theoretical Domain Framework (TDF) approaches, is a guide to the structured analysis of behaviours and provides a framework to support the development of behaviour change interventions.

The BCW COM-B/TDF approaches were selected for use in the study following a short scoping review of behaviour change frameworks (see discussion section below), and used to help design topic guides and the development of the Training & Communication Package. The BCW (incorporating COM-B & TDF) was chosen over the other frameworks because of its breadth across environment and contextual factors, and the wide-ranging review and consolidation process underpinning the TDF [17].

The BCW has been used in two ways. Firstly, key behavioural categories were identified from the TDF and reflected as prompts in the topic guide template (later adapted by the study sites). Secondly, in developing the T&C package, the BCW was used to prompt discussions on possible intervention categories by site research teams.

### Iterative, collaborative and adaptive approaches

The research methods and approach were developed in an iterative and collaborative process between the core study team and the 8 research partner organisations across the 5 countries in the study. Building on collaborative protocol design workshops in Geneva and Delhi, discussions online and site visits, core methods and templates were developed.

Local research teams adapted core methods and templates based on local context, populations, cultural and research norms. The level of adaption was designed to ensure that the research methods and tools would be as appropriate as possible for the local context, resulting in locally appropriate results, within the scope of a common and overarching protocol.

Core methods include:

1. Use of qualitative methods, including use of content analysis
2. Purposive sampling
3. Flexible and adjustable In-depth Interviews and Focus Group Discussion (FGD) guides
4. Audio recording, transcribing and translating of the interviews and discussions
5. Use of 2 independent coders and a 3rd external coder to resolve coding conflicts
6. Manual or software-based coding
7. Preliminary and final analysis

To ensure rigor and increase authenticity in our methodology, we used triangulation through two independent coder and a third independent coder to resolve coding conflicts.

Topic guide templates for In-Depth Interviews (IDIs) and Focus Group Discussion (FGDs) were developed based on the research questions, with the design of prompts influenced by selection of relevant TDF categories from the BCW.

The topic guides include factors that help, or hinder:

1. the effective communication of healthcare workers with patients/caregivers about adherence to the prescription
2. obtaining prescribed medicine by patients and caregivers and ensuring the adult patient or child takes the medicine correctly following the prescription instructions

Topic guide prompts cover the TDF areas of:

1. Knowledge
2. Skills
3. Belief about capabilities
4. Memory, attention and decision processes
5. Physical opportunity
6. Social influences/professional identify
7. Belief about consequences
8. Routines and habits

Purposive sampling is used to identify participants for the qualitative study, who are enrolled after providing written consent. Purposive sampling is widely used to identify and select information-rich participants for the most effective use of limited resources [18]. Individuals are selected who are especially knowledgeable about the topic of interest [19], in this case adult patients and caregivers in the community local to the study clinics. Participants must be available, willing to participate and have the ability to communicate experiences and opinions in an articulate, expressive, and reflective manner [20].

Confidentiality was ensured throughout the research process to protect participants from harm [21, 22]. Locations for FGDs and IDIs were selected to provide confidentiality in data gathering, all personal identifiable information was removed from participant responses in the process of transcribing audio recordings, and from any accompanying notes; in drafting reports we are careful to ensure that any participant responses, included to illustrate research findings, are not identifiable by personal characteristics.

While there are many approaches to saturation [23]; we are influenced by the description of Code and Meaning saturation [24, 25], though there is a degree of flexibility and difference in the choice of approach to saturation across the studies in the different sites. Data collection responses are regularly reviewed to consider levels of saturation and to guide expansion or halting data collection. At a minimum code saturation is to be reached, while the level of meaning saturation will vary by code and research site [24].

Content analysis is used to develop a code framework from the transcripts, followed by application of the framework to code meaning units in the transcripts, coded manually or in software, by two coders, with a third coder checking and resolving differences. Analysis builds on meaning units to develop categories and themes as described by Erlingsson et al [26].

### Site adaption

Local research teams in the five countries adapted the methods and topic guide templates outlined above in the areas of:

- Participant groups:
  for healthcare workers participant groups varies depending on the staff roles within the study health clinics;
  for potential patients and caregivers of the clinic, this varies based on the characteristics of the population around the study clinics.
- Matching of methods to different participant groups
- Locations for data collection
- Compensation for participants
- Language used for data collection, transcriptions, analysis and reporting

In all the study research sites there are a number of different local and national languages that could be used at the different stages in the research process. To determine the language(s) to be used in each site we reviewed language use at the following stages:

1. Data gathering – IDI and FGDs,
2. Audio recording and any additional notes taken,
3. Transcription of audio recording,
4. Development of codes and analysis,
5. Development of communication messages and training content
6. Delivery of training
7. Delivery of communication messages during the trial,
8. Publication of research methods and results.

Overall, the level of adaption was designed to ensure that the application of research methods in each site would be as appropriate as possible and build on the local research team’s experience and knowledge of local populations and cultural norms.

### Patient and public involvement

Patients and the public more generally were not involved in the development of the research methods described here.

### Methods to Develop the T&C package

The Training and Communication (T&C) Package consists of communication messages for patients and caregivers, and training for healthcare workers in delivery of the messages of adherence to the prescription. The T&C package is implemented in two stages, healthcare workers are trained in communicating messages of prescription adherence during the clinical trial site initiation, and the messages of adherence are communicated to patients and caregivers as part of the intervention package for patients in the intervention arm of the study, during initial (Day 0) consultations.

The contents of the T&C package are developed to respond to the behaviour determinants identified in the research analysis. Specifically, Behaviour determinants are considered to be the research Themes identified in the Context Analysis process. Themes emerge as the highest level of abstraction from the research analysis, building on categories and codes, grouping together meaning from across one or more categories in the data [26].

Consequences for the T&C package of each behaviour determinant are identified considering:

- the content of the message,
- how the messages will be delivered,
- how to train the healthcare workers in communicating the message,
- or the behaviour determinant is identified as unable to be influenced through the T&C package (e.g. antibiotic stock levels).

Each Theme/behaviour determinant in the research, is considered separately, before the consequences are combined to form the T&C package - either in the communication messages, including tools such as infographics, or in training plans. Some adaption may take place to make sense of overlapping factors, when forming the T&C package as a whole.

To support this process, the BCW was used to identify possible intervention categories, triggering discussion and debate on a range of responses to the behaviour determinants, by the study teams.

### SARS-CoV-2

Site research teams are currently exploring how to adapt the use of the research methodologies described here to adhere to physical distancing measures needed in SARS-CoV-2 prevention and response.

## Discussion

### Integration of qualitative research with a clinical trial

Qualitative research can potentially add value to clinical trials in the areas of bias, efficiency, ethics, implementation, interpretation, relevance, success and validity [12], but in doing so tensions are created which should be addressed.

In developing the research protocols, there can be a tension and at times confusion between the two possible aims of the study – to answer social science questions, and to develop a social science element that complements a quantitative component of the trial. The iterative protocol development process discussed these two approaches before clarity emerged that the focus of the qualitative work was to support and complement a quantitative (clinical) study. In this study, the clinical trial approach was prioritised; in a pure social science study, we might expect the effects of the delivery of prescription adherence messages to be evaluated by comparing adherence before and after the delivery of the messages. In contrast, in the randomised control trial approach implemented in this study, the effect of delivery of T&C messages of adherence to prescription is evaluated through the comparison of adherence in the intervention and control arms (collected through a qualitative discussion on Day 7), in keeping with the overall analysis approach in the trial.

However, because the T&C intervention is incorporated into a package of interventions for the trial, we cannot rule out the influence of other intervention components, for example, in the use of an increased number of diagnostics, on prescription adherence. Similarly, due to the intervention package approach, we cannot conclude how much the T&C package contributed to the primary outcome (patient recovered at Day7). However, notwithstanding these limitations, the approach is appropriate as a deductive extension of relatively proven methods. Given that few behaviour change interventions will be generalizable across locations, the process highlights an approach to rapid, local context appropriate delivery support.

### Selection of behaviour change framework

For simplicity a single behaviour change framework was chosen to underpin the behavioural approach taken in the study. We selected the Behaviour Change Wheel (BCW) guide [6, 7], incorporating the Capacity, Opportunity and Motivation (COM-B) and Theoretical Domains Framework (TDF) following a small scoping review of behavioural frameworks. The scoping review was conducted considering the BEHAVE framework [27], Precede-Proceed model [28]; Motivation, Ability, Prompt (MAP) model [29]; Normalised Process Theory [30]; Health Belief Model [31, 32] and the Behaviour Change Wheel. The COM-B and TDF approach, as described in the BCW guide, was considered the most appropriate to capture the wide range of potential social and cultural issues that may drive prescription adherence and diagnostic use behaviours, and for its discussion of intervention functions driven from behavioural determinants.

### Language considerations

As discussed above, we considered the language(s) to be used in multiple stages in the data gathering, analysis and reporting process. In making these choices we considered that:

1. greater levels of translation result in more opportunities to introduce errors,
2. translation and the required back translation for quality control, takes precious time.

It is clear that to obtain the best quality data the language used for in-depth-interviews and focus group discussions must be is the first language of the participant, and if that is not possible, then a second language that is understood and spoken well by both the participant and researchers.

In general, when choosing the languages for use in other stages, we prioritised fewer translations, meaning higher quality with fewer errors, resulting in a faster process, over the need for documents to be available in English (or French) for international audiences, and applied this prioritisation from an early stage in the process.

### Linking research findings to T&C package

The two subject of qualitative research methods, and behavioural approaches, contain their own process, concepts and terminologies. In using the research findings to develop the T&C package for the clinical trial, we have bridged the gap between the language of Content Analysis and behaviour analysis used in the BCW. This has been implemented by equating Themes that emerge from content analysis, which is the analysis of discussions on behaviours, to behaviour determinants used for the BCW. Our experience using this approach, has been positive and all key findings from the research have translated across into the behavioural approach.

### Limitations

The approach chosen is not without its limitations:

Firstly, the intervention is limited to messages of adherence that can be delivered within the patients/caregivers’ visit to the clinic at the time of diagnosis and prescription. It does not allow for a wide range of other interventions which may support adherence to prescriptions, such as repeated messages of adherence to the patient while they are completing the prescription. Further, the scope of the intervention does not allow for any community wide influencing strategies, as such strategies may impact on community members who go on to be study participants in the control arm, who must not receive the intervention.

Within the scope of the study, further activities could usefully include extensive testing of variations in communication messages and expanding participation in the process to develop the T&C package. Both of these steps would strengthen the T&C package.

Overall, and from a behavioural perspective, we expect communication of messages on adherence to prescription from the healthcare workers to the patients and caregivers to be only one part of a wider strategy that is needed to support prescription adherence behaviours in the community.

## Data Availability

The datasets used and/or analysed during the study are availablefrom the corresponding author upon reasonable request.

## Acknowledgements

We would like to thank the qualitative research teams in Ghana, Uganda, Burkina Faso, Nepal, Myanmar, and India who have contributed to local adaptation of study methods and research methods. Additional thanks to Heidi Hopkins, London School of Hygiene and Tropical Medicine, UK, for her support.

## Ethical approval

The work described in this paper has received ethical approval as part of the approval of the overall trial protocol by the Oxford University clinical trials research ethics committee (OxTREC number 52-19). Each of the country-specific protocols is also approved by national and/or institutional ethics committees in Burkina Faso, Ghana, Uganda, India and Nepal.

After being informed of the study details, potential participants must provide written consent before they can engage further in the study.

